# Performance of ChatGPT on Clinical Medicine Entrance Examination for Chinese Postgraduate in Chinese

**DOI:** 10.1101/2023.04.12.23288452

**Authors:** Xiao Liu, Changchang Fang, Ziwei Yan, Xiaoling Liu, Yuan Jiang, Zhengyu Cao, Maoxiong Wu, Zhiteng Chen, Jianyong Ma, Peng Yu, Wengen Zhu, Ayiguli Abudukeremu, Yue Wang, Yangxin Chen, Yuling Zhang, Jingfeng Wang

**Author notes:** **Corresponding author:** Yuling Zhang, Jingfeng Wang. Co-first author (Xiao Liu, Changchang Fang).

## Abstract

**Background:** The ChatGPT, a Large-scale language models-based Artificial intelligence (AI), has fueled interest in medical care. However, the ability of AI to understand and generate text is constrained by the quality and quantity of training data available for that language. This study aims to provide qualitative feedback on ChatGPT’s problem-solving capabilities in medical education and clinical decisionmaking in Chinese.

**Methods:** A dataset of Clinical Medicine Entrance Examination for Chinese Postgraduate was used to assess the effectiveness of ChatGPT3.5 in medical knowledge in Chinese language. The indictor of accuracy, concordance (explaining affirms the answer) and frequency of insights was used to assess performance of ChatGPT in original and encoding medical questions.

**Result:** According to our evaluation, ChatGPT received a score of 153.5/300 for original questions in Chinese, which is slightly above the passing threshold of 129/300. Additionally, ChatGPT showed low accuracy in answering open-ended medical questions, with total accuracy of 31.5%. While ChatGPT demonstrated a commendable level of concordance (achieving 90% concordance across all questions) and generated innovative insights for most problems (at least one significant insight for 80% of all questions).

**Conclusion:** ChatGPT’s performance was suboptimal for medical education and clinical decision-making in Chinese compared with in English. However, ChatGPT demonstrated high internal concordance and generated multiple insights in Chinese language. Further research should investigate language-based differences in ChatGPT’s healthcare performance.

## Introduction

Artificial intelligence (AI) is initially conceptualized in 1956^1^, but it has only gained significant momentum in recent years. AI aims to replicate human intelligence and thinking processes through the use of brain-like computer systems to solve complex problems. The most inspiring is that, AI systems can be trained on specific data sets to improve prediction accuracy and address intricate problems ^2-4^, which means that one of the possible application of AI is the ability to helps doctors to rapidly search through vast amounts of medical data, enhancing their creativity and enabling them to make error-free decisions^5, 6^.

ChatGPT is an artificial intelligence model that has spurred great attention due to the revolutionary innovations in its ability to perform a diverse array of natural language tasks. By using a class of Large-scale language models (LLMs), GPT3.5 can predict the likelihood of a sequence of words based on the context of the preceding words. With sufficient training on vast amounts of text data, ChatGPT can generate novel word sequences that closely resemble natural human language, but have never been observed before by other AI^7^.

A study was conducted on the effectiveness of the version of GPT LLM (GPT3.5) in passing the United States Medical Licensing Exam (USMLE). The results showed that the AI model achieved an accuracy rate of over 50% in all the tests, and in some analyses, it even surpassed 60% accuracy. It is imperative to highlight and emphasize that the study was conducted mostly using English input and the AI model was also trained in English.

However, like all language models, ChatGPT’s ability to understand and generate text in any given language is limited by the quality and quantity of training data available in that language. Chinese is the second most widely spoken language in the world, with more than 1.3 billion speakers globally, while the quality and quantity of Chinese language data may be not compared with English due to some reasons, such as complexity of the written characters. Thus, the performance of ChatGPT in Chinese medical information warrants further investigation.

In this study, we evaluate ChatGPT’s clinical reasoning ability by administering questions from the Clinical Medicine Entrance Examination for Chinese Postgraduate in Chinese. This standardized and regulated test assesses candidates’ comprehensive abilities, the questions textually and conceptually dense, and the difficulty and complexity of questions is highly standardized and regulated. Additionally, this exam has demonstrated remarkable stability in raw scores and psychometric properties over the past years. Moreover, the exam comprises 43% basic science and medical humanities, with 14% physiology, 10% biochemistry, 13% pathology, and 6% medical humanities. Clinical medicine makes up the remaining 57%, with internal medicine and surgery accounting for 37% and 20%, respectively. Due to the exam’s linguistic and conceptual complexity, we hypothesize that it will serve as an excellent challenge for ChatGPT.

## Methods

### Artificial Intelligence

ChatGPT is a state-of-the-art language model that employs self-attention mechanisms and vast training data to produce natural language responses in a conversational context. Its key strengths include the ability to handle long-range dependencies and generate coherent, contextually relevant replies. However, it is worth noting that GPT3.5 is a server-based language model that lacks internet browsing and search capabilities. As a result, all responses generated are based solely on the abstract relationships between words, or “tokens,” within its neural network^7^. It should be noticed is that the OpenAI has developed a latest version GPT4 in March 2023, while the inputting date is Feb 2023 which the latest version is GPT3.5 at that time.

### Input source

The test questions for the Chinese Clinical Medicine Postgraduate Entrance Examination in 2022 is not released by the official website. However, a complete set of 165 questions with a total of 500 point was available online (Supplemental S1), which is deemed as original questions. The point values for questions varied. Case analysis questions were worth 2 points each, as were the Multi-Choice questions. Additionally, there were 60 Common Questions worth 1.5 points each, and 30 other Common Questions worth 2 points each.

All the inputs given for the GPT-3.5 model are valid samples that do not belong to the training dataset. This is because the database has not been updated since September 2021, which is prior to the release of these questions. In order to streamline future research efforts, the 165 questions have been grouped into three distinct categories, as listed below.

1. Common Questions (n=90): These questions are to evaluate the knowledge in basic science in physiology, biochemistry, pathology, and medical humanities. There are four choices for each question, and the respondent should collect the only correct answers. For example: “The closing time of the aortic valve during the cardiac cycle is? A. Atrial systolic end card B. Rapid ejection beginning C. Slow ejection beginning D. Isovolumic diastole beginning”
2. Case Analysis Questions (n=45). It is a method used in clinical medicine to examine and evaluate patient cases. It involves an in-depth review of a patient’s medical history, presenting symptoms, laboratory and imaging results, and diagnostic findings to arrive at a diagnosis and treatment plan. There are four choices, and the respondent should collect the only correct answers. The difference between Case Analysis Questions and Common Questions is that Common Questions is focus on clinical decision making. For example: “A 38-year-old male, suffering chest pain and fever for 3 days, having a 5 years of diabetes history. Physical examination: T=37.6LJ, right lower lung turbid knock, breathing sound is reduced. A chest X radiograph suggests a right pleural effusion. Pleural aspiration liquefaction test showed WBC650×10^6^/L with fine lymph Cell 90% in pleural fluid, with glucose of 3.2 mmol/L, the diagnosis for this patient is? A. Tuberculous pleurisy B. malignant pleural effusion C. empyema D. pneumonia-like pleural effusion”
3. Multi-Choices Questions (n=30): There are four choices, and the respondent should collect the correct answers which is more than two. There are no points for choosing more or less. For example: “The structures of auditory bone conduction include? A. skull B. round window film C. ossicular chain D. cochlear bone wall”.

### Scoring

Initially, we realized that modifying the question format was necessary to accurately evaluate ChatGPT’s performance in questions of the Chinese Clinical Medicine Postgraduate Entrance Examination. Specifically, we found that reminding the AI with “multi-choice” or “single-choice” was essential as ChatGPT produced varying results without this specification. For the Multi-Choices Questions, we modified it to read “Please choose one or more of the correct options,” while the Common Questions and Case Analysis Questions were modified to read “There is only one correct answer.” This only applies when evaluating the score of ChatGPT in answering questions in the Chinese language.

We created a dataset consisting of questions from the Chinese Clinical Medicine Postgraduate Entrance Examination and their corresponding answers. To ensure its accuracy, we verified these answers by comparing them with those available on the internet and consulting with senior doctors. We then used this dataset to evaluate ChatGPT’s performance on the exam by comparing its responses to the standard answer. A high score on the exam would indicate that ChatGPT performed well on this task.

### Encoding

To better reflect the actual clinical situation, we modified these questions to be open-ended. We presented the Case Analysis Questions to ChatGPT in different variations without multiple-choice options and asked it to identify the disease the patient had and explain its reasoning. For the Multi-Choices Questions, we removed all the choices without reminding ChatGPT that there were multiple options. For the Common Questions, we processed them in the same manner as the Multi-Choices Questions. However, there was an exclusive group within the three subgroups that could not be encoded in the same way as the others. These questions required selecting one of the provided choices, and thus, we transformed them into a special form (n=26), which is highlighted in yellow in Supplemental S1. For example, the original question, “Which can inhibit insulin secretion? A. Increased free fatty acids in blood B. Increased gastric inhibitory peptide secretion C. Sympathetic nerve excitation D. Growth hormone secretion increases” was encoded as “Can an increase in free fatty acids in the blood, an increase in gastric inhibitory peptide, an increase in sympathetic nerve excitation, or an increase in growth hormone secretion can inhibit insulin secretion?” The encoder was present in all three subgroups.

Furthermore, to minimize memory retention bias, a new chat session was initiated for every enquiry.

### Adjudication

AI outputs were independently scored for Accuracy, Concordance, and Insight by two physician who were blinded to each other, adjudicators using the pre-defined criteria Supplemental S2. A subset of 20 questions was used to train the physician adjudicators who were not blinded to each other. The accuracy of ChatGPT’s responses was classified into three categories: accurate, inaccurate, and indeterminate.

Accurate responses mean that ChatGPT provided the correct answer. Inaccurate responses included no answer, an incorrect answer, or multiple answers with incorrect options. Indeterminate responses imply that the AI output does not provide a definitive answer selection, or it believes that there is insufficient information to do so. Concordance was defined as the ChatGPT’s explanation affirms its provided answer, while a discordant explanation contradicts it. Valuable insights were defined as unique instances of text within the AI’s explanations that met specific criteria: they were nondefinitional, nonobvious, valid, and unique. Specifically, valuable insights required additional knowledge or deduction beyond the input question, provided accurate clinical or numerical information, and had the potential to eliminate multiple answer choices with a single insight.

To reduce within-item anchoring bias, the adjudicators evaluated accuracy for all items first, followed by concordance for all items. Two physicians were blinded to each other. If there was discrepancy on the domains, a third physician adjudicator was consulted. The number of third adjudicator for Common Questions and Multi-Choices Questions was 7 and 3, respectively. The need for third adjudicator in Case Analysis Questions was 1 for concordance. Ultimately, 11 items (6.8% of the dataset) required the intervention of a third physician adjudicator. The interrater agreement between the physicians was evaluated using the Cohen kappa (κ) statistic for the questions (Supplemental S3).

A schematic overview of the study protocol is provided in Fig 1.

**Fig 1.**
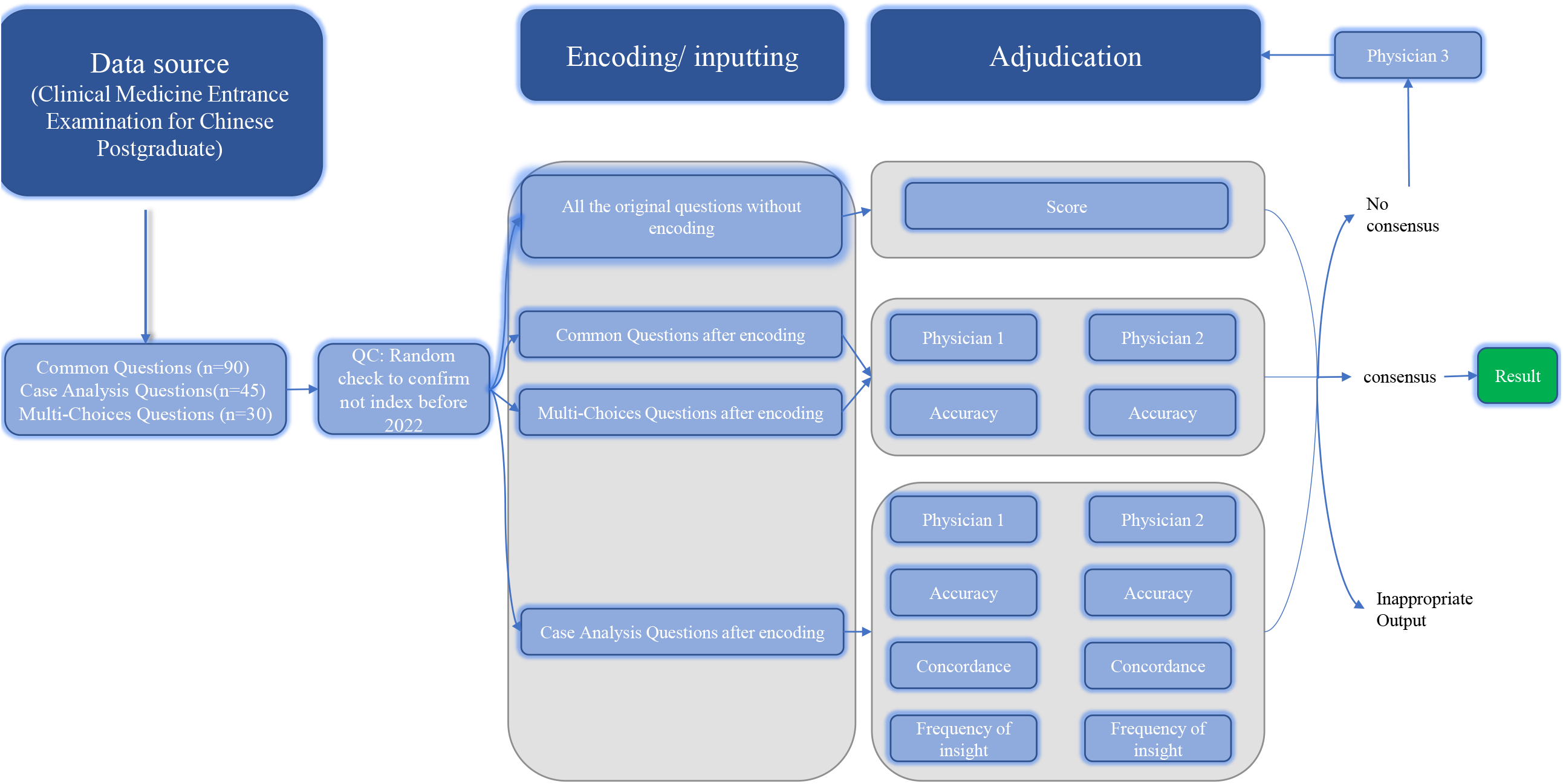
Schematic of workflow for sourcing, encoding, and adjudicating results. Abbreviations: **CQ** =Common Questions; **CAQ** =Case Analysis Questions; **MCQ** =Multi-Choices Questions. The 165 questions were categorized into three types: CQ, CAQ, and MCQ, and each question was assessed for its score. The accuracy of the CQ and MCQ questions were evaluated, while the MCQ questions were also assessed for the accuracy, concordance, and frequency of insights. The adjudication process was carried out by two physicians, and in case of any discrepancies in the domains, a third physician was consulted for adjudication. Additionally, any inappropriate output was identified and required re-encoding.

## Result

### ChatGPT performance poor towards the original questions

After inputting the original questions into ChatGPT and collecting its answers, ChatGPT received a score of 153.5/300, which means that it only obtained 51.16% of the total points on the test. This score is much lower than the expectation, but slightly higher than the passing threshold (129/300) defined by official agencies.

Among three subgroups of questions, the evaluation revealed that out of a total of 90 Common Questions, ChatGPT only provided 50 (55.6%) correct answers. Similarly, out of 45 Case Analysis Questions, ChatGPT provided 25 (55.6%) correct answers. Furthermore, out of 30 Multi-Choices Questions, ChatGPT provided 10 (33.3%) completely accurate answers (Fig 2). These results suggest that ChatGPT’s ability to resolve medical problems in Chinese needs to be improvement.

**Fig 2.**
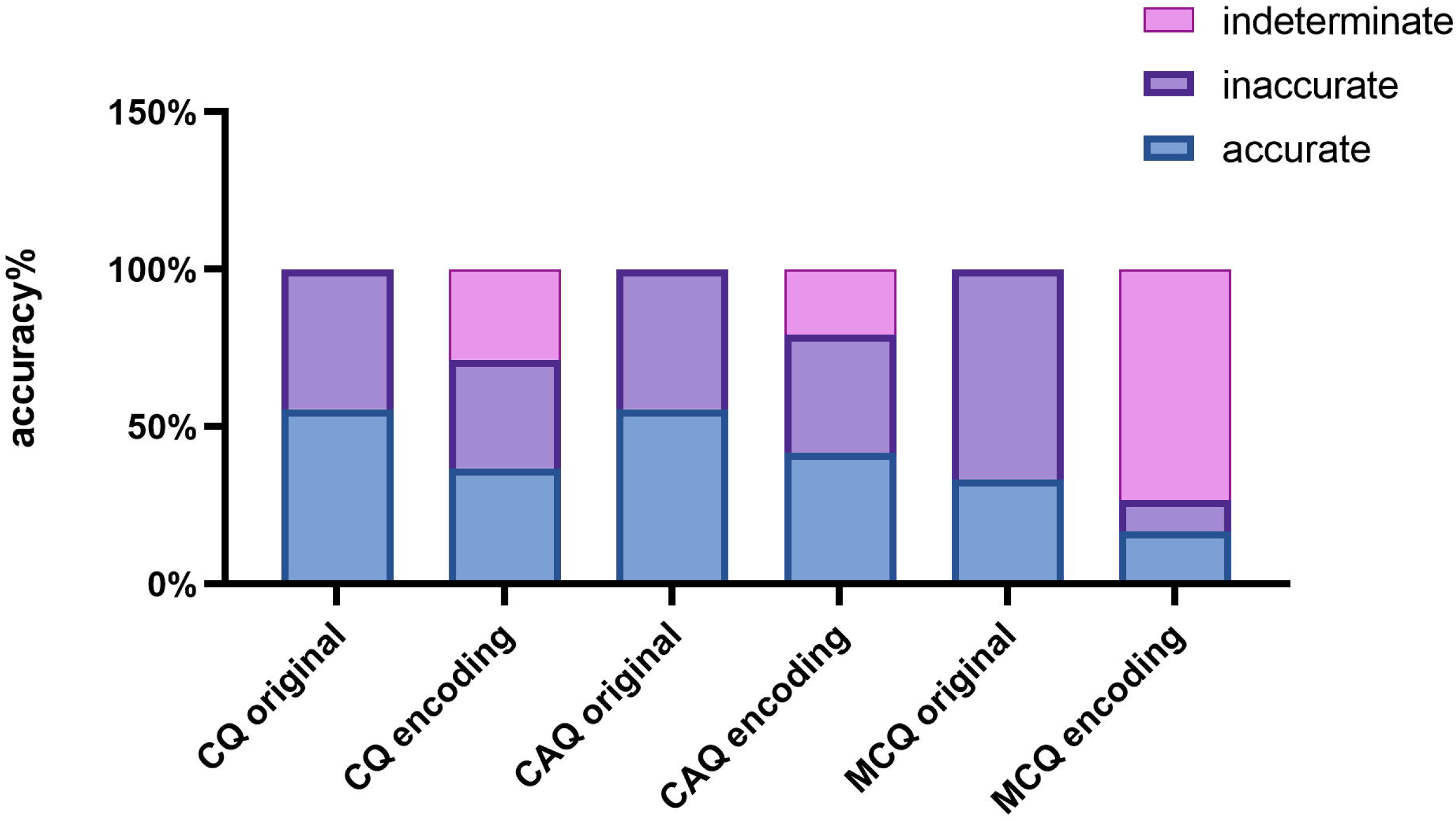
Accuracy of ChatGPT on Chinese Clinical Medicine Postgraduate Entrance Examination Test before and after encoding. For the subgroup CQ, CAQ and Multi-Choices Questions before encoding, AI output were compring with the standard answer key. For the subgroup CQ, CAQ and Multi-Choices Questions after encoding, AI outputs were adjudicated to be accurate, inaccurate, or indeterminate based on the scoring system provided in S2 Data. It demonstrates the different accuracy distribution for inputs between the before and the after.

### ChatGPT performs worse on encoded questions compared to the original questions

We encoded questions of the Chinese Clinical Medicine Postgraduate Entrance Examination and inputted them into ChatGPT, which simulates scenarios where a student asks a common medical question without answer choices, or a doctor tries to diagnose a patient based on multimodal clinical data (i.e. symptoms, history, physical examination, laboratory values). ChatGPT’s accuracy was 31.5% for all questions.

Among the three subgroups, namely Common Questions, Multi-Choices Questions, and Case Analysis Questions, the accuracy was 41.7%, 36.8%, and 16.7%, respectively (Fig 2). Compared the original questions, the accuracy of their encoding questions decreased by was 19%, 17%, and 14%, for Common Questions, MultiChoices Questions, and Case Analysis Questions, respectively, which demonstrates the ability of ChatGPT answering the open-ended questions in Chines is shortcoming. During the adjudication stage, there was substantial agreement among physicians for prompts in all three subgroups (κ ranged from 0.80 to 1.00).

#### ChatGPT demonstrates high internal concordance

Concordance, which is a measure of the level of agreement or similarity between the option selected by AI and its subsequent explanation, was also taken into consideration. The results showed that ChatGPT had 90% concordance across all questions, and this high concordance was maintained across all three subgroups (Fig 3). Additionally, we analyzed the concordance difference between correct and incorrect answers and found that concordance among accurate responses was perfect and significantly greater than among inaccurate responses (100% vs. 50%, p<0.001) (Fig 3). These findings suggest that ChatGPT has a high level of answer-explanation concordance in Chinese, likely due to its strong internal consistency in its probabilistic language model.

**Fig 3.**
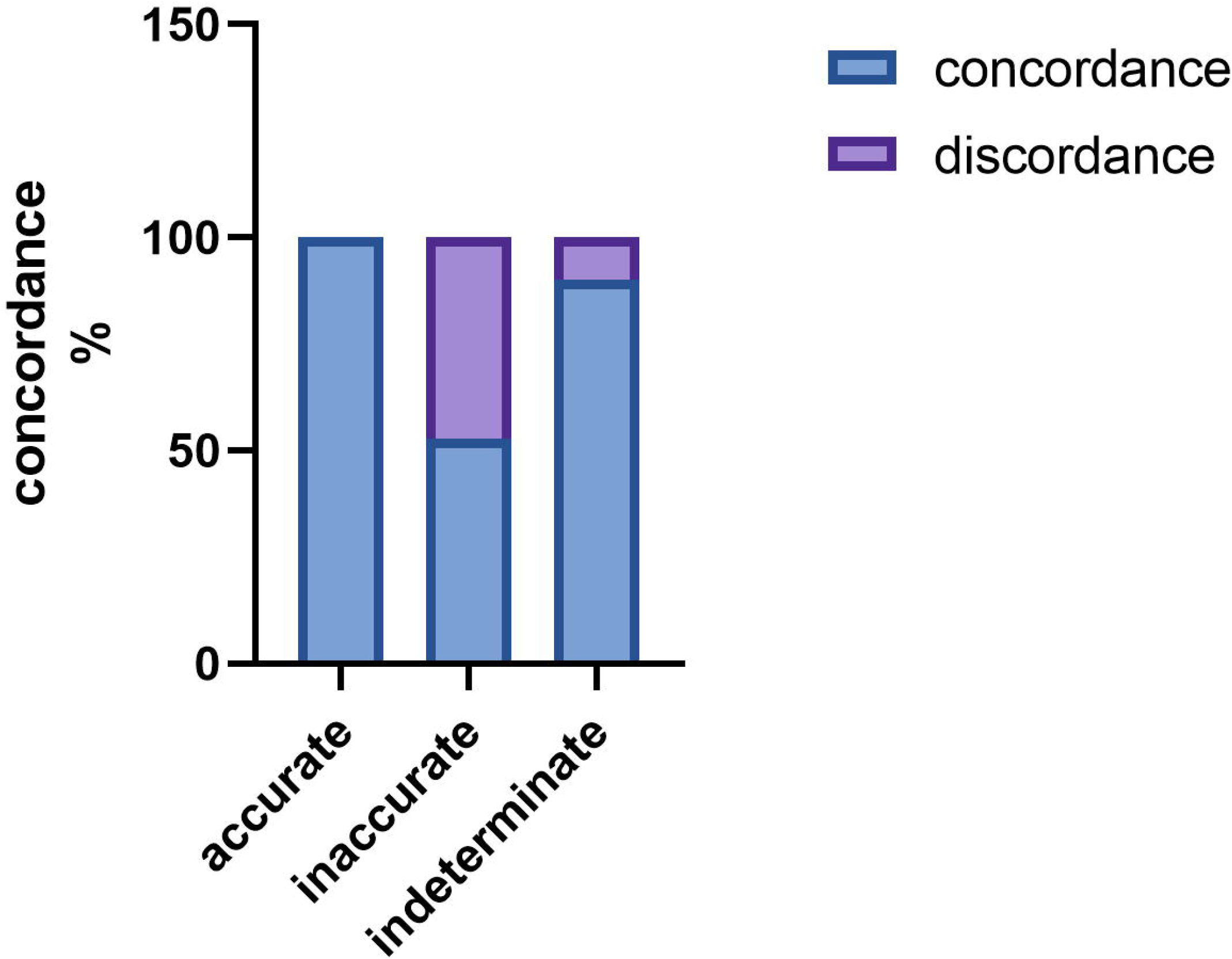
Concordance of ChatGPT on Chinese Clinical Medicine Postgraduate Entrance Examination after encoding. For the subgroup “case analysis question”, AI outputs were adjudicated to be concordant and discordant, based on the scoring system provided in S2 Data. It demonstrates concordance rates stratified between accurate, inaccurate and indeterminate outputs, across all of the CAQ

#### ChatGPT shows multiply insights towards the same questions

Another evaluation index considered was the frequency of insights generated by the AI model, which quantifies the quantity of insights produced. After evaluating the score, accuracy, and concordance of ChatGPT, we investigated its potential to enhance medical education by augmenting human learning. We examined the frequency of insights provided by ChatGPT. Remarkably, ChatGPT generated at least one significant insight in 80% of all questions (Fig 4). Moreover, the analysis revealed that the accuracy response had the highest frequency of insights, with an average of 2.95. The indeterminate response followed closely behind with an average of 2.7, while the inaccurate response had a lower frequency of insights with an average of 1.39 (Fig 4). The high frequency of insights in the accurate group suggests that it may be feasible for a target learner to acquire new or remedial knowledge from the ChatGPT AI output.

**Fig 4.**
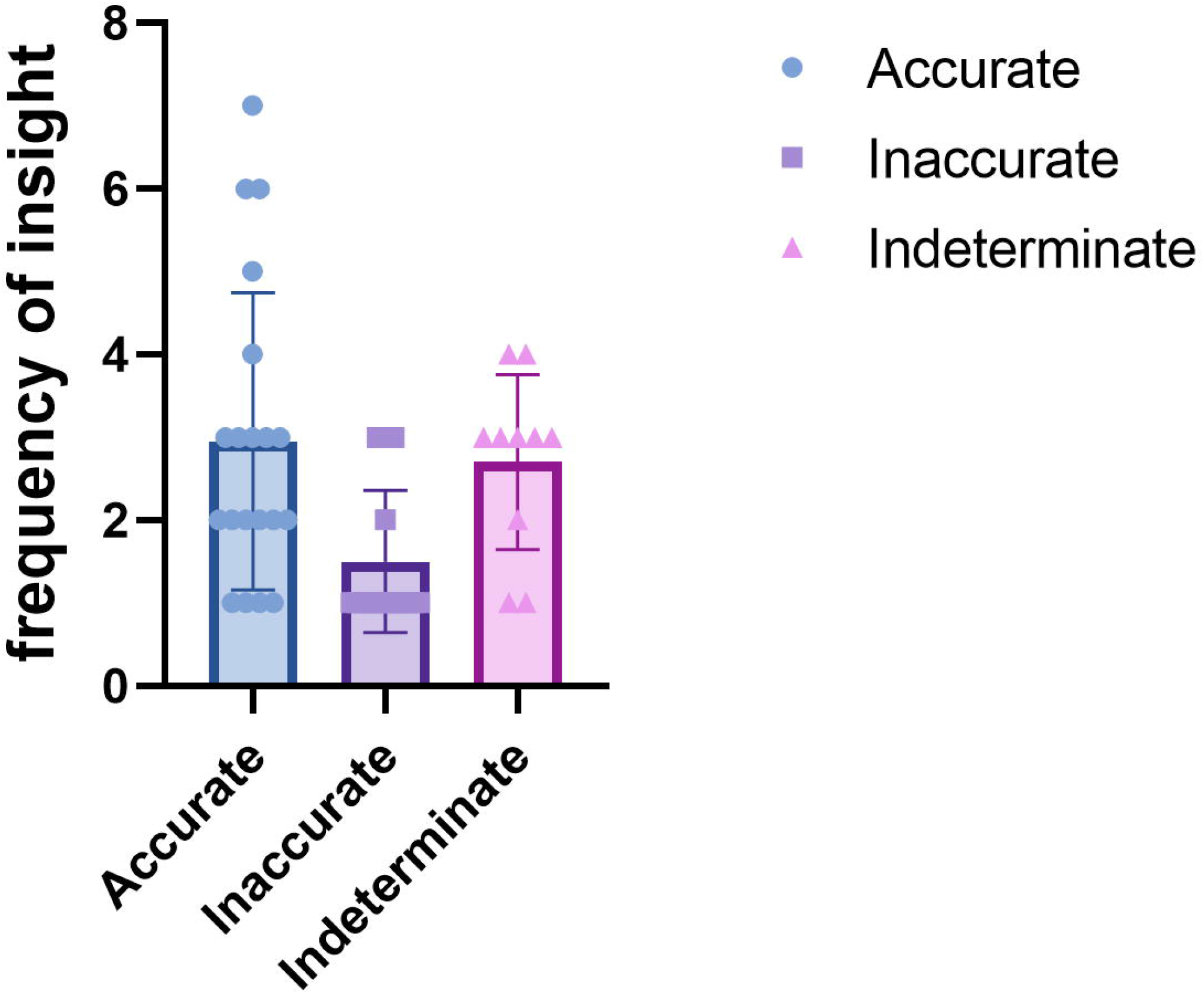
The frequency of insights of ChatGPT on Chinese Clinical Medicine Postgraduate Entrance Examination after encoding. For the subgroup “case analysis question”, AI outputs were adjudicated to count the frequency of insights it offered. It demonstrates frequency of insights stratified between accurate, inaccurate and indeterminate outputs, across all of the CAQ.

## Discussion

### Major findings

To evaluate ChatGPT’s problem-solving capabilities and assess its potential for integration into medical education in Chinese, we tested its performance on the Chinese Clinical Medicine Postgraduate Entrance Examination. Our findings can be organized into 2 major themes: (1) The score of ChatGPT, which needs to be improved when facing questions asking in Chinese language; (2) There are still potential for this AI to generate novel performance that can assist human due to the high concordance and the frequency of insights. This is the first study to assess the performance of ChatGPT on in medical care and clinical decision in Chinese.

### ChatGPT performance need improvement for medical questions in Chinese

A recent study showed the ChatGPT3.5 performed with an accuracy rate of over 50% across all examinations and even exceeded 60% accuracy in some analyses when facing the United States Medical Licensing Exam (USMLE)^7^.In our results, the study found that ChatGPT exhibited moderate accuracy in answering open-ended medical questions in Chinese, with accuracy was 31.5%. Given the differences between English and Chinese inputs, we conclude that ChatGPT requires further improvement in answering medical questions in the Chinese language.

We sought to understand why there is a significant discrepancy between the performance of ChatGPT on Chinese and English language exams. To investigate this, we asked the ChatGPT for the reason, it explains that the training data used to train AI in different languages may be different, and the algorithms used to process and analyze text may vary from language to language (data not shown). Therefore, even for the same question, the output generated may vary slightly based on the language and the available language-based data.

Upon analyzing the results of our research, we found that the accuracy of ChatGPT was lowest for Multi-Choices Questions, followed by Common Questions, and Case Analysis Questions. The lower accuracy on Multi-Choices Questions s may be due to the model being undertrained on the input, as well as the Multi-Choices Questions samples being significantly less than those of single-choice questions. On the other hand, the Case Analysis Questions may have extensive training compared to Multi-Choices Questions, is similar in type to the USMLE question.

Furthermore, we noticed that high accuracy outputs were associated with high concordance and a high frequency of insight, whereas poorer accuracy was linked to lower concordance and a lack of insight. Thus, we hypothesized that inaccurate responses were primarily driven by missing information, which could result in reduced insight and indecision in the AI, rather than an over-commitment to an incorrect answer^7^. The results indicate that enhancing the database and providing additional training with Chinese questions could lead to a substantial improvement in the performance of the model.

### Challenges of AI in future applications

Despite the promising potential of AI in medicine, it also poses some challenges. Standards for use of AI in health care are still need to be developed^8, 9^, including clinical care, quality, safety, malpractice, and communication guidelines. Furthermore, the implementation of AI in healthcare requires a shift in medical culture, which poses a challenge for both medical education and practice. Additionally, ethical considerations must be taken into account, such as data privacy, informed consent, and bias prevention, to ensure that AI is used ethically and for the benefit of patients. Surprisingly, A recently launched AI system for autonomous detection of diabetic retinopathy carries medical malpractice and liability insurance^10^.

### Prospective of AI

AI is a rapidly growing technology. At this time of writing, the ChatGPT has released version 4 with great improvement. Numerous practical and observational studies have demonstrated the versatile role of AI in almost all medical disciplines and specialties, particularly in improving risk assessment,^11, 12^data reduction, clinical decision support^13, 14^, operational efficiency, and patient communication^15, 16^. We anticipate that advanced language models such as ChatGPT are reaching a level of maturity that will soon have a significant impact on clinical medicine, enhancing the delivery of personalized, compassionate, and scalable healthcare.

### Limitations

With the limitation of our research is the small sample size. We only access 165 samples to qualify its accuracy and 30 case analysis questions to qualify its concordance and frequency of insight, Furthermore, the clinical situation is more complicated than the test, larger and deep analyses were needed. Finally, bias and error were inevitable for human-adjudication, although there was a good interrater agreement between the physicians for the adjudication.

## Conclusion

In conclusion, although the ChatGPT’s got a score over the passing score in Clinical Medicine Entrance Examination for Chinese Postgraduate in Chinese language, the performance was limited when presented with open-ended questions. On the other hand, ChatGPT demonstrated a high level of internal concordance, which suggests that the explanations provided by ChatGPT support and affirm the given answers. Moreover, ChatGPT generated multiple insights towards the same questions, demonstrating its potential for generating a variety of useful information. Further prospective studies are needed to explore whether there was a language-based difference in performance of medical education setting and clinical decision-making, such as Chinese and minority language.

## Data Availability

All data produced in the present study are available upon reasonable request to the authors
All data produced in the present work are contained in the manuscript

## Data statement

All data generated or analyzed during this study are included in this published article [and its supplementary information files].

## Acknowledgments

We acknowledge the ChatGPT for polishing our manuscript.

## Author contributions

X-L and W.G.-Z was responsible for the entire project and revised the draft. J. W-C, K.B-M and Q. W-H performed the study selection, data extraction, statistical analysis, and interpretation of the data. J. W-C and X.L. drafted the first version of the manuscript. All authors participated in the interpretation of the results and prepared the final version of the manuscript.

## Declarations

Ethics approval This is a systematic review and meta-analysis. No ethical approval is required.

