## supplemental table for "Performance of ChatGPT on Clinical Medicine Entrance Examination for Chinese Postgraduate in Chinese"

### Data Supplement

Table S1： The original questions

The raw data file can be accessed at the following URL:

https://drive.google.com/file/d/17_1mGKbN5MLoK6yZmSL363Q1wjIJB3wO/view?usp=share_link

Table S2： Adjudication criteria for accuracy, concordance.

| | |
|---|---|
| Accurate:1. Provide the answer accurately.<br>2.When the judge determines that there is not a unique answer, the AI outputs multiple choices, among which contains the correct answer, and the other options are also completely correct. | Concordant: Explaining affirms the answer |
| Inaccurate 1. No answer is provided<br>2.An incorrect answer is provided<br>3.Multiple answers are provided, among which there is an incorrect answer, even if the correct answer is included. | Discordant: Explanations are contradictory. |
| Indeterminate: 1.AI output is not a single answer election.<br>2.When the judge determines that there is a unique answer, AI output provides multiple choices.<br>3.AI believes that there is not enough information." | |

TableS3：Kappa statistic for interrater agreement between adjudicating physicians.

| | Accuracy | | Concordance | |
|---|---|---|---|---|
| | Cohen κ | n | Cohen κ | n |
| CAQ | 0.559 | 45 | 0.444 | 45 |
| MCQ | 0.795 | 30 | | |
| CQ | 0.825 | 90 | | |
